# Plasma cell-free DNA methylome as early detection and prognostic marker for pancreatic cancer

**DOI:** 10.1101/2025.05.29.25328569

**Authors:** Sanjeev Budhathoki, Yonathan Brhane, Gordon Fehringer, Shu Yi Shen, Dianne Chadwick, Philip C. Zuzarte, Ayelet Borgida, Daniel D. De Carvalho, Steven Gallinger, Rayjean J. Hung

## Abstract

Pancreatic cancer is highly fatal, with limited early detection options. Here, we conducted a study to evaluate the potential use of circulating cell-free DNA methylation for pancreatic cancer early detection and prognosis. Using a highly sensitive enrichment-based sequencing technology, we profiled genome-wide circulating cell-free methylome of 199 pancreatic cancer patients and 205 healthy individuals. We identified a panel of differentially methylated regions in cell-free DNA that distinguished pancreatic cancer cases from controls with high accuracy (test set AUC=0.93, 95%CI=0.88-0.98), notably robust accuracy for early-stage pancreatic cancer (AUC=0.92, 95%CI=0.86-0.98). We also identified a panel of cfDNA methylated regions that effectively stratified patients into longer or shorter survival groups, which was independently validated in The Cancer Genome Atlas cohort (HR=1.37, 95%CI=1.00-1.88, p=0.049). These results provide support for the potential utility of cell-free DNA methylation markers for both early detection and prognosis in pancreatic cancer, presenting a promising tool for enhancing patient management.

## INTRODUCTION

Pancreatic cancer remains one of the most fatal cancers, with about 90% of patients dying within 1 year of diagnosis^1^. The majority of pancreatic cancer patients are diagnosed at an advanced stage, where prognosis is poor with a 5-year survival rate of approximately 5%^2^. Early detection of pancreatic cancer is critical, as evidenced by the more favorable prognosis with a 5-year survival of about 80% for patients with resectable tumors in stage 1^2^. However, few patients are diagnosed in these earliest stages, underscoring the need for identification of sensitive and accurate biomarkers for early detection^2, 3^.

Epigenetic alterations, including DNA methylation, have a pivotal role in carcinogenesis^4^. For pancreatic cancer in particular, the importance of epigenetic alterations is highlighted by aberrant methylation of cancer related genes such as CDKN2A, RASSF1A, SERPINB5 and S100P in pancreatic tumour tissue ^5^. Furthermore, comprehensive multi-omics analysis combining methylation profiling and RNA expression data in The Cancer Genome Atlas (TCGA) study has identified genes that are recurrently silenced by DNA methylation in pancreatic ductal adenocarcinoma ^6^. Aberrant DNA methylation patterns occur early in tumorigenesis, making them attractive targets for diagnostic and prognostic biomarkers of pancreatic cancer. The continuously improved technologies enabling the sensitive detection of methylated cell-free DNA (cfDNA) fragments in the circulation offers a minimally invasive approach for detection and monitoring pancreatic cancer development^7, 8^. Recently, leveraging circulating cfDNA methylation markers, multicancer early detection tests are being developed to detect multiple cancer types and evaluated in cancer screening setting ^9, 10, 11^.

Specifically for pancreatic cancer, several studies have evaluated the utility of cfDNA methylation markers in early detection and prognosis of pancreatic cancer. However, these previous studies typically investigated a small number of targeted methylation regions and reported low sensitivity to detect pancreatic cancer ^8, 12^. Moreover, most of the previous studies had small sample sizes (N=30-90) and lacked sufficient statistical power to reliably determine the potential for early detection or prognosis. Genome-wide profiling of DNA methylation in cell-free DNA offers a comprehensive and agnostic approach to investigate differential methylation profiles, but it is challenging due to the low amount of DNA available and the small fragment size of circulating DNA, generally less than 200bp in length. We have developed a novel and sensitive method to interrogate the genome-wide methylation profile, and using this method we demonstrated that the cell-free methylome can be used to trace back to the tumor of origin with high accuracy ^7^. In the current study, we conducted an in-depth analysis focused on pancreatic cancer based on the largest pancreatic cancer dataset generated to date, with the goal to validate the utility of cell-free DNA methylome for pancreatic cancer early detection, and its potential utility as a prognostic marker.

## RESULTS

### A genome-wide analysis of cell-free DNA methylome in pancreatic cancer cases and healthy controls

Based on 199 pancreatic cancer cases and 205 healthy controls frequency matched on age and sex, we conducted genome-wide methylation profiles using cell-free methylated DNA immunoprecipitation sequencing (cfMeDIP-seq), as previously described ^7, 13^. The key characteristics of pancreatic cancer patients and controls are shown in **Table 1**. As expected, pancreatic cancer patients had higher body mass index, more tobacco smoking history, and higher percentage family history than controls.

**Table 1.** Distribution of selected characteristics of the pancreatic cancer cases and controls.

|  | Cases (n=199) |  | Controls (n=205) |  |
| --- | --- | --- | --- | --- |
|  | No. | (%) | No. | (%) |
| Sex |  |  |  |  |
| Men | 101 | (50.7) | 103 | (50.2) |
| Women | 98 | (49.2) | 102 | (49.8) |
| Age*, mean (SD), years | 66.6 | (11.2) | 66.8 | (11.2) |
| BMI*, mean (SD), Kg/m<sup>2</sup> | 27.6 | (4.5) | 25.8 | (4.9) |
| Smoking Status |  |  |  |  |
| No | 90 | (48.4) | 104 | (51.5) |
| Yes | 96 | (51.6) | 98 | (48.5) |
| Missing | 13 |  | 3 |  |
| Pack years of smoking*, mean (SD), | 15.3 | (20.5) | 8.4 | (14.5) |
| Family history of pancreatic cancer |  |  |  |  |
| No | 171 | (88.6) | 196 | (95.6) |
| Yes | 22 | (11.4) | 9 | (4.4) |
| Missing | 6 |  |  |  |
| Stage |  |  |  |  |
| I-II | 112 | (56.3) |  |  |
| III-IV | 85 | (42.7) |  |  |
| Unknown | 2 |  |  |  |

\*Values are means and standard deviation (SD)

For the purpose of model training and testing, we split the study sample into 70% training set for model development and 30% test set for model performance evaluation. Methylation data were obtained from a total of 8,946,446 regions. Based on the negative-binomial model on training set, we identified 151,703 differentially methylated regions (DMRs) in the pancreatic cancer cell-free methylome profile compared to healthy controls (Benjamin-Hochberg adjusted *p-value* <0.05; **Supplementary Figure 1A**).

To assess whether the DMRs detected using cfMeDIP-seq reflect the methylation aberrations in tumour tissue samples from this study population, we compared the plasma cell-free methylome to somatic methylation profiles generated from 24 matched tumour-normal tissue samples from a subset of the study participants using reduced representation bisulfite sequencing ^7^. A total of 4,675 differentially methylated cytosines (DMCs) found in paired tumour normal tissue analysis were detected in cell-free DNA of the same individuals (**Supplementary Figure 1B)**. Based on the permutation analysis, we observed highly significant concordance between the cell-free methylome and tumour tissue methylation patterns (concordantly hypermethylated *P*= <5×10^−1000^ and concordantly hypomethylated *P* = 8.6×10^−136^) (**Supplementary Figure 1C**).

### Cell-free methylation profiles and prediction of pancreatic cancer status

We next evaluated how well cell-free methylation profiles can be used to discriminate pancreatic cases from controls. In the training set, we fitted a model using the 4,675 DMRs that were concordant with tissue methylation profiles from the tumour-normal pair comparison described above. Using the elastic-net penalty function and 5-fold cross validation, we identified 217 DMRs that best predicted case-control status in the training set after accounting for marker correlations **(Supplementary Table 1)**. We then computed the cell-free Methylation Risk Score (cfMRS) using penalized logistic regression analysis with these methylation markers. Plotting the individual cfMRS values in the test set by key patient characteristics showed that the only factor that is clearly clustering in two ends of the cfMRS distribution is pancreatic cancer status (**Figure 1A**). No clear pattern with other key patient characteristics was observed, which means that cfMRS distribution not driven by other patient factors. **Figure 1B** shows that in the test set, the distribution of the cfMRS for pancreatic cancer patients is substantially higher compared to healthy controls. Based on the model developed using the training set, we then estimated the area under the receiver operating characteristics curve (AUC) to assess the added value of the methylation markers, based on models with the cell-free methylation marker alone, with known risk factors of pancreatic cancer including cigarette smoking status, body mass index (BMI), family history of cancer and age and sex, and in combination with both cfMRS and risk factors. In the test set, the model containing risk factors only conferred an AUC of 0.73 (95%CI=0.63-0.83). The predictive performance improved substantially to 0.93 (95%CI=0.88-0.98) after adding the cfMRS (**Figure 1C**). Using the cutoff value based on Youden index method, the sensitivity and specificity of the covariate and cfMRS combined model was 0.85 and 0.91 respectively.

**Figure 1.**
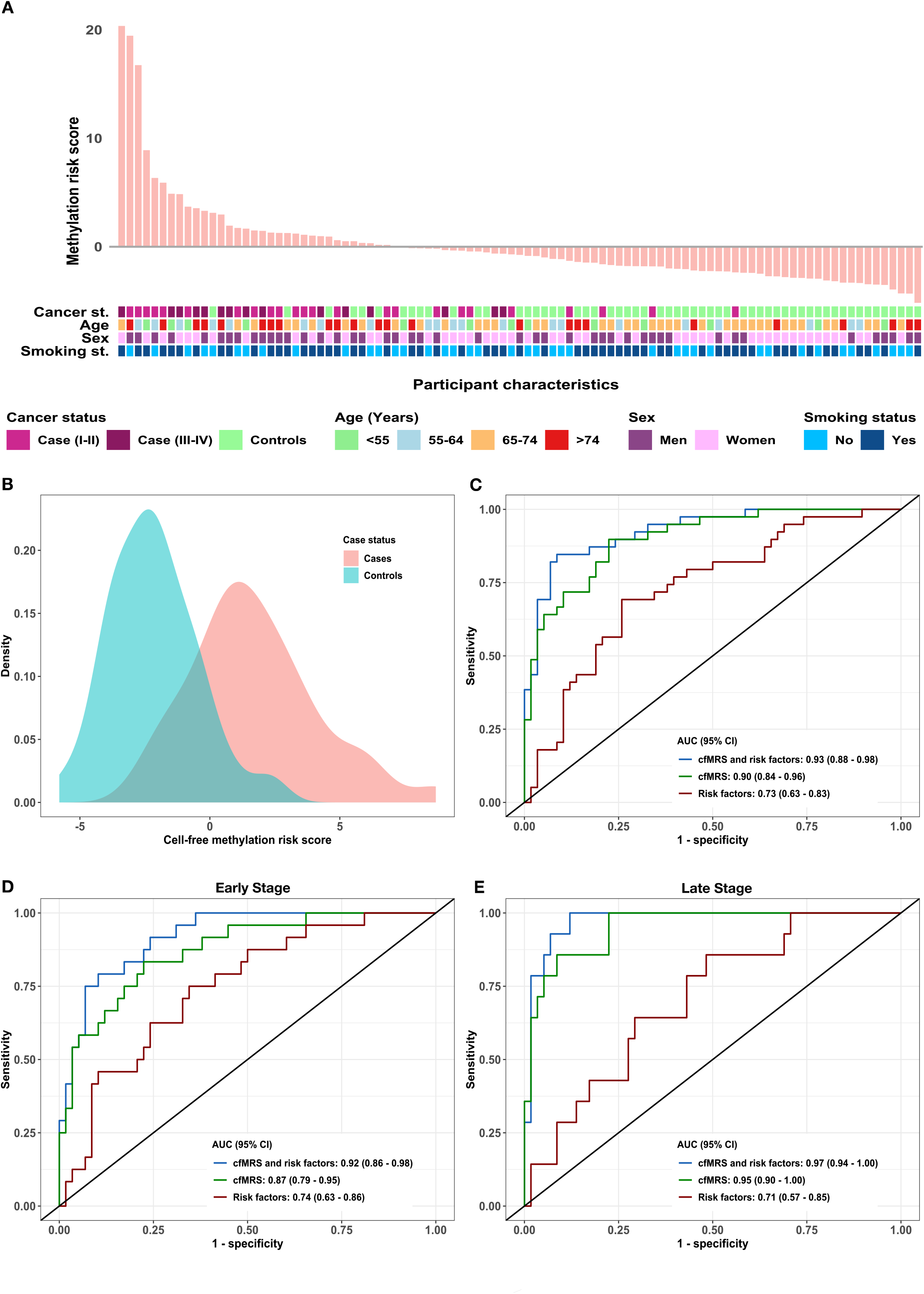
Cell-free methylation profiles and prediction of pancreatic cancer presence in the test set. **(A)** Distribution of cell-free methylation risk score by selected characteristics in the test set. Y-axis represents the cell-free methylation risk scores based on the differential methylated regions, and X-axis represent each individual in the test set (N=114). Different patient characteristics are annotated per legends-including cancer status (non-cancer controls marked in green, pancreatic cancer cases are in different shades of magenta according to their stage at diagnosis), age (<55, 55-64, 65-74, >74 years), sex (Men vs Women) and smoking status (No vs Yes). **(B)** Distribution of the cell-free methylation risk score (cfMRS) among pancreatic adenocarcinoma cases and controls. **(C)** ROC curve discriminating pancreatic cancer cases and controls in test set based on 3 different models: risk factors only, cfMRS only, and combining both risk factors and cfMRS for overall dataset. **(D)** ROC curve discriminating pancreatic cancer cases and controls in test set based on 3 different models in early stages; and **(E)** ROC curve discriminating pancreatic cancer cases and controls in late stage. Risk factors model includes age, sex, body mass index, smoking status, history of diabetes, and family history of pancreatic cancer.

To assess the model performance for early-stage pancreatic cancer, which is most relevant for early detection, we stratified the test set into early (stage I and II) and late stage (stage III and IV). For early stage pancreatic cancer, model predictive performance is highest when combining cfMRS and risk factors (AUC=0.92, 95%CI=0.86-0.98), as opposed to a model based on risk factors only (AUC=0.74, 95%CI=0.63-0.86) (**Figure 1D**). The results suggest that the methylation markers are effective in identifying early-stage pancreatic cancer patients. The AUC based on cfMRS and risk factors was higher in late-stage cancer (AUC=0.97, 95%CI=0.94-1.00) (**Figure 1E**). This is expected since more tumor DNA is shed into the circulatory system in late stages. The sensitivity and specificity of the cfMRS-risk factor combined model was 0.79 and 0.90 respectively for early stage cancer, and 0.99 and 0.88 respectively for late stage cancer. Notably the cell-free methylation risk score alone reached overall AUC of 0.90 (95%CI=0.84-0.96) (**Figure 1C**), and the discriminatory performance remained good when stratified into early-stage (AUC=0.87, 95%CI=0.79-0.95) (**Figure 1D**) and late-stage cancer (AUC=0.95, 95%CI=0.90-1.00) **(Figure 1E)**.

We further performed overrepresentation enrichment analyses using Gene Ontology (biological process) terms to evaluate the biological pathways that are enriched for 217 DMRs found to be most predictive for the presence of pancreatic cancer. This analysis showed the enrichment of terms related to cell development, differentiation and multicellular organism development (**Figure 2A**). The network analysis revealed 5 major components of the network that are separated into 2 clusters: one with 4 linked components (regulation of metabolic process, establishment of localization, signal transduction, cellular component organization or biogenesis), and one as a single cluster (cell adhesion) (**Figure 2B**).

**Figure 2.**
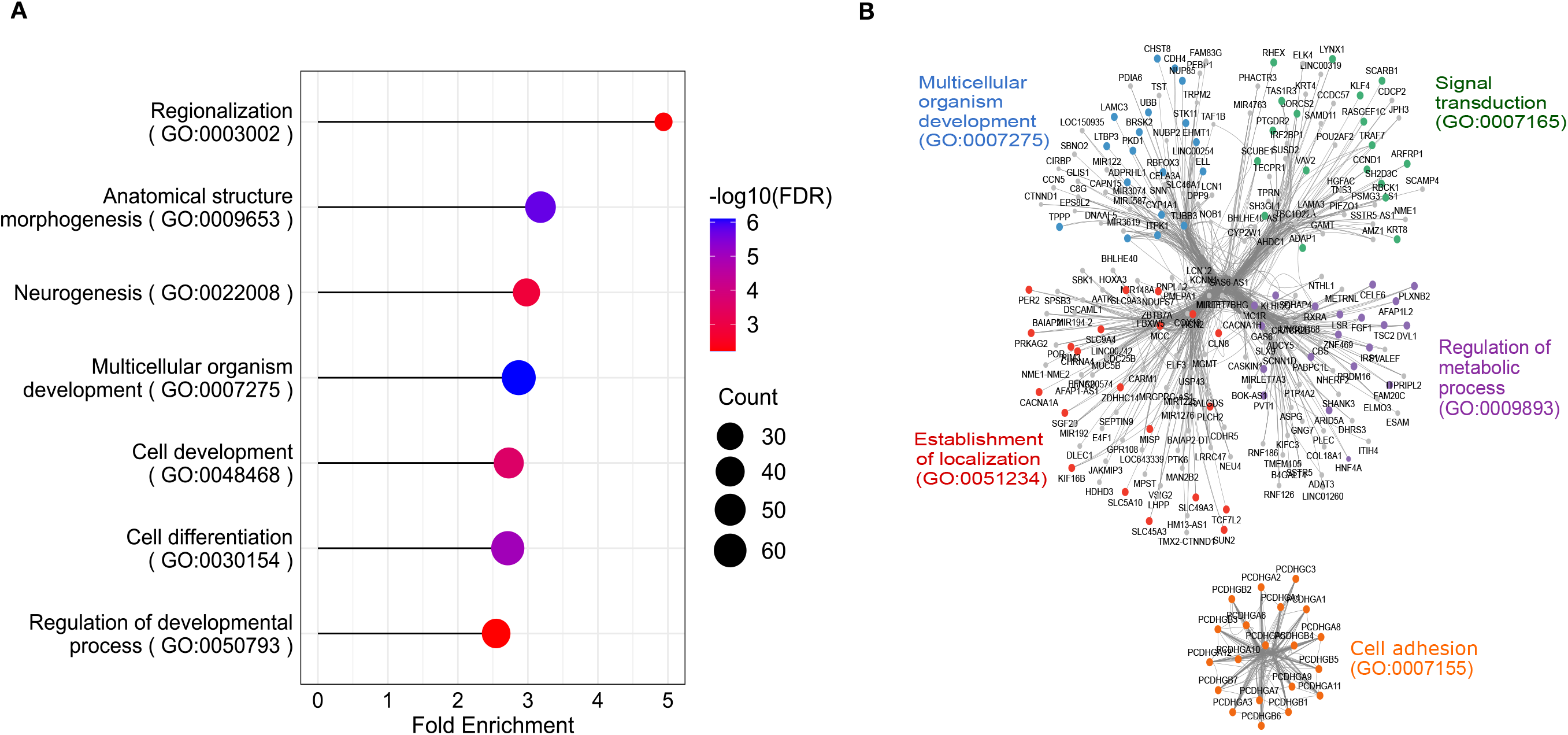
Pathway and network analysis for differential methylated regions that indicate the presence of pancreatic cancer. **(A)** Gene ontology (GO) enrichment analysis (biological process terms), FDR, false discovery rate; Count, number of overlapping genes in each pathway. **(B)** weighted correlation network analysis of methylated DMRs. Each node represents a gene and line represents the connection between genes based on strength of co-expression. Genes that were identified to be in the same network are depicted in clusters.

### Cell-free methylation profiles and prediction of survival

Using the cell-free methylome profile of 199 pancreatic cancer patients, we evaluated the prognostic value of methylation markers for prediction of survival, after accounting for clinical factors. To identify the differential methylation regions associated with survival and to reduce the dimensionality of the methylation profiles, we first assessed each DMR in a Cox-regression model adjusted for age and sex. A total of 6003 methylation regions were retained at a Benjamini-Hochberg adjusted significance level of p<0.05. We then performed penalized regression analysis with Lasso penalty parameter in the retained markers based on the nested cross-validation method. The methylation hazard scores were computed as the sum of aberrant DMRs weighted by corresponding hazard ratios after penalized regression. A total of 48 DMRs had a non-zero coefficient and contributed to the methylation hazard score (MHS). We divided pancreatic cancer patients into high- and low-hazard groups based on the median of the methylation hazard score. Plotting individual methylation hazard score distribution showed higher score in advanced stage cases and lower score in early-stage cases (**Figure 3A**), and the MHS is not driven by any other key patient characteristics, such as age, sex, and smoking status. It also showed negative correlation with survival (**Figure 3B** and **3C**). The log-rank test showed that high-MHS group had significantly much poor survival (median=285 days, 95% CI= 255-317 days) than the low-MHS group (median=915 days, 95% CI= 730-1208 days) **(Figure 3D**) (log rank P: <0.001). The effect of other key prognostic factors are shown in **Supplemental Figure 2**, and as expected, stage is one of the main prognostic factors. In a multivariable analysis, the methylation hazard score was significantly associated with higher probability of death and remained an independent predictor of shorter survival after accounting for age, sex and cancer stage, with an adjusted hazard ratio (HR) of 4.79 (95%CI=3.24-7.08).

**Figure 3.**
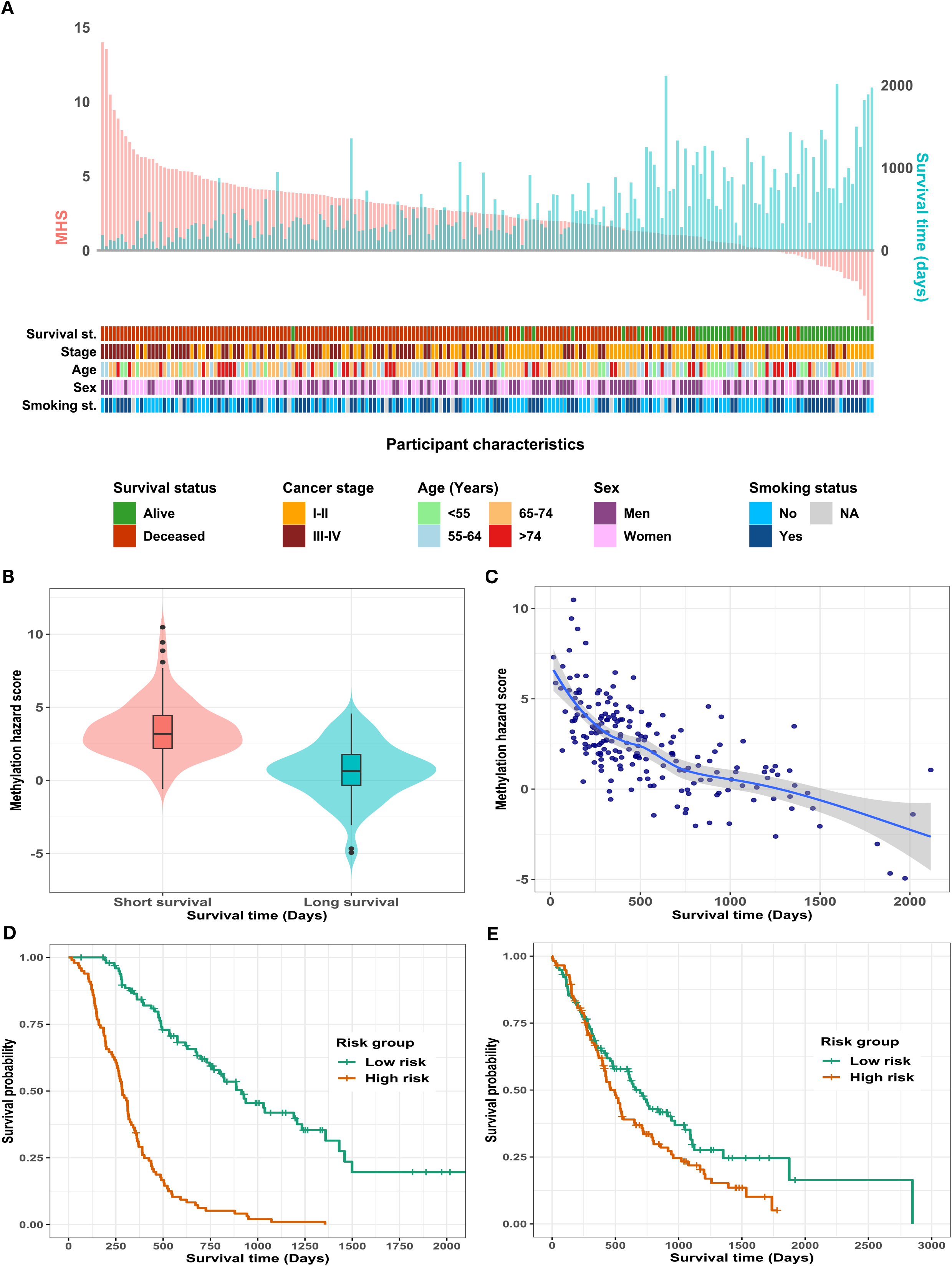
Cell free methylation profiles and prediction of prognosis and survival. **(A)** Distribution of cell-free methylation hazard score in the pancreatic cancer cases by selected characteristics. Y-axis represents the individual cell-free methylation hazard scores (peach colour) and survival time (blue colour), and X-axis represent each pancreatic cancer cases (N=199). Different patient characteristics are annotated per legends-including survival status (Alive vs Deceased), cancer stage (I-II vs III-IV), age (<55, 55-64, 65-74, >74 years), sex (Men vs Women) and smoking status (No vs Yes). **(B)** Distribution of methylation hazard score in cancer cases by median survival time. **(C)** Relationship between methylation hazard score and survival time. **(D)** Kaplan-Meier survival curve of pancreatic cancer by methylation hazard score categories. **(E)** Kaplan-Meier survival curve of pancreatic cancer by risk score categories in validation (TCGA/ICGC) data.

### Validation of prognostic markers in TCGA and ICGC cohort

To externally validate the prognostic value of the cell-free methylation markers, we evaluated the prognostic potential of these methylation markers based on TCGA and ICGC data. We identified the methylation sites in TCGA and ICGC Infinium Methylation Array that overlapped the methylation regions in our data and followed the same analysis steps as above. Sixteen out of 48 DMRs were matched in the TCGA and ICGC dataset. Consistent with the result we observed in our data set, patients with a high methylation hazard score had a shorter survival time than those with a low methylation hazard score (**Figure 3E**). The median survival stratified by methylation hazard score was 695 (95%CI=480-913) days in the low-hazard group and 495 (95%CI=400-552) days in the high-hazard group (Log rank test P value <0.05), with HR of 1.37 (95%CI=1.00-1.88).

## DISCUSSION

This is the largest study to date of the cell-free methylome and pancreatic cancer. Strengthened by matched tumor data in a subset of study, we identified a panel of cell-free methylation markers that could reliably classify the presence of pancreatic cancer with high accuracy, even at an early-stage, indicating that cfDNA methylation assays may serve as a tool to detect early-stage pancreatic cancer. In addition, our results showed that methylation markers can stratify patients into those with longer and shorter survival times in both our study and independently in TCGA/ICGC, suggesting utility of cell-free DNA methylation markers in pancreatic cancer prognosis.

We employed a genome-wide approach to identify the methylation alteration sites which is an advantage of our study compared to array technologies that cover a small portion of (1% to 5%) the methylation sites in the genome^14, 15^. This led to the identification of a large number of differentially methylated regions that are available to distinguish pancreatic cancer patients from healthy individuals, which helped to increase the sensitivity, specificity and prediction accuracy.

Identifying pancreatic cancer at an early stage is important for improving prognosis since survival is poor for locally advanced or metastatic cases. However, detection of methylation patterns in circulating cell-free DNA is particularly challenging for early-stage tumors with lower tumor burden. Aided by our genome-wide approach and highly sensitive method, we observed comparable prediction accuracy, sensitivity and specificity for both late and early-stage cancers indicating that our methylation marker panel may be suitable as an early detection assay. Early detection of pancreatic cancer can lead to survival advantages. Albeit currently no established screening program exist for pancreatic cancer, intensive surveillance among high-risk individuals has shown to lead to smaller tumor at diagnosis^16^. It is conceivable to complement the identification of high risk individuals based on family history with the liquid biopsy.

In recent years, multi-cancer early detection (MCED) studies based on cell-free methylome have shown the potential use of methylation markers in early detection of cancer, although the sample sizes for pancreatic cancer tend to be very limited. Lui et al. tested methylation markers for multi-cancer early detection that included 84 pancreatic cancer cases in the training and 39 in the validation set) ^10^, and reported sensitivity of 63% for pancreatic cancer stage I (N=8 in validation set). Another multicancer study in NHS hospital sites (including 12 pancreatic cancer patients), reported overall sensitivity of 91.7% for pancreatic cancer at specificity of 98.4% ^11^. However, in this study the accuracy is based on inclusion of up to two predicted cancer signal origins, which likely inflated both sensitivity and specificity because it artificially improves the chances of correctly identifying cancers. This can pose challenges for subsequent clinical management^9^. In addition, both above studies had limited numbers of pancreatic cancer cases compared to non-cases and this imbalance between cancer cases and controls can bias the model toward higher specificity but lower sensitivity. This is particularly relevant for rare cancers like pancreatic cancer because the case/control imbalance may cause the model to perform less reliably on cancers with fewer cases, as the model could overfit to the control (non-cancer) samples.

Employing a prospective study would minimize some of the biases in the above studies. Furthermore, prospective cohorts allow for the observation of temporal relationships between DNA methylation patterns and the subsequent development of pancreatic cancer, providing a better understanding of causality and aiding in the identification of early biomarkers for detection and prevention. Upon proper validation in prospective cohorts, we consider that MCED and cancer-specific assays may be complementary, depending on the tumor types. For relatively rare but fatal cancer such as pancreatic cancer, it is particularly crucial to estimate the assay performance accurately specific to the tumor type to optimize the subsequent patient management.

Regarding the prognostic value of cell free DNA markers, some of the prior investigations used the presence of circulating tumor DNA before or after surgery, or before and after adjuvant therapies, as a marker to predict the prognosis of pancreatic cancer, while others focussed on recurrent somatic mutations in the circulating tumor DNA^17, 18^. However, circulating tumor DNA tends to have low sensitivity, especially for early-stage cancer, and therefore their prognostic utility is less reliable as seen from the inconsistent results from these studies^17^. Some other studies used methylation markers derived from tumour tissue sequencing to predict prognosis of pancreatic cancer^19, 20^. In our analysis, we found that specific cell-free methylation markers are an independent prognostic factor in predicting pancreatic cancer survival and they can stratify pancreatic cancer cases with long versus poor survival group. Moreover, using the methylation markers identified in our study, we were also able to replicate the prognostic potential of the methylation markers in the TCGA and ICGC cohort, albeit the association with survival was less marked compared to ours, which is likely due to the difference in the techniques (cfMeDIP-seq in our dataset vs Illumina Infinium Human Methylation 450 BeadChip Array in TCGA/ICGC) used to obtain methylation profiles. Identifying patients with high risk and low probability of death within a defined time period can help to optimize treatment strategies.

In summary, we have identified a specific panel of cell-free methylation markers that can accurately identify the presence of pancreatic cancer including early-stage diseases, and a panel of cell-free methylation markers that can predict pancreatic cancer survival. Our findings indicate that cell-free DNA methylation markers hold significant potential as biomarkers for early detection of pancreatic cancer and prediction of pancreatic cancer prognosis. Although there is growing attention to multi-cancer early detection panels, sensitivity and specificity of the multi-cancer approach remains a concern, particularly related to false positive results. A combination of methylation markers with other molecular and clinical factors could enhance diagnostic and prognostic models, enabling more precise risk stratification. Future studies should focus on validating these findings in large prospective cohorts and exploring the potential of combining methylation markers with other molecular data to further refine diagnostic and prognostic tools for pancreatic cancer.

## METHODS

### Study samples

Pancreatic cancer patients and healthy controls were recruited as part of the Ontario Pancreas Cancer Study^21^ and Multicancer case-control study in Ontario^22^. Briefly, eligible cases were residents of Ontario with a first primary, pathologically confirmed exocrine tumour of the pancreas. Controls were healthy individuals recruited through the Family Medicine Centre at Mount Sinai Hospital and frequency matched to cases on age and sex. Both cases and controls completed self-administered questionnaires regarding family history, medical history and lifestyle risk factors and clinical data were available for all cases. Plasma samples were collected from all controls and from all cases before treatment. In addition, tumour and adjacent normal tissue samples were obtained for 24 cancer patients. All subjects provided written informed consent to participate in the study. All samples were obtained upon approval of the institutional ethics committees and Research Ethics Boards from Mount Sinai Hospital, in compliance with all relevant ethical regulations.

### Specimen Processing for patient cfDNA

Plasma samples from cases and controls were collected using EDTA and acid citrate dextrose tubes and stored at −80°C until assayed. Cell-free DNA was extracted from 0.5-3.5 ml of plasma using the QIAamp Circulating Nucleic Acid Kit (Qiagen). The extracted DNA was quantified through Qubit prior to use^7^.

### Methylation profiles and data processing

The genome wide methylation profiles of pancreatic cancer cases and matched controls from plasma cfDNA were generated using cell-free Methylated DNA Immunoprecipitation and high-throughput sequencing method (cfMeDIP-seq). This method was specifically designed for the cfDNA methylome analysis to overcome the low amount and fragmented nature of cfDNA and is described in detail previously^7, 13^.

The raw cfMeDIP-seq data was aligned to the human genome (version hg19) using Bowtie. The SAM files obtained were converted to BAM files using Samtools and input into the MEDIPS package for downstream bioinformatics analysis. The sequencing reads were binned to 300 bp non-overlapping methylated regions (windows) and coverage profiles (reads per kilobase per million mapped reads (RPKMS)) were then obtained for each window^7, 13^.

### Statistical analysis

#### Differentially methylated region (DMR) analysis and classification of disease status

We filtered out windows with less than 10 read counts across all samples. The read count for each window was then normalized by trimmed mean of m-values (TMM) normalization method. The varying library sizes were adjusted for using quantile-adjusted conditional maximum likelihood method. Subsequently, after fitting negative binomial model and dispersion estimation, the exact test was used for testing the differential methylation region. DMR analysis was done using the R package *edgeR*^23^.

We collected pancreatic tumour and normal tissue samples from 24 patients^7^. Genome-wide methylation profiles from the extracted DNA of these tumour and normal tissue samples were created using Reduced Representation Bisulfite Sequencing (RRBS). We had identified 45,372 differentially methylated cytosines (DMCs) from the tumour-normal tissue sample analysis. We performed concordance analysis between DMRs identified from cfDNA and the DMCs from tumour-normal tissue sample.

We divided cases and controls into two groups: a training set (70%) to develop the model, and a test set (30%) to validate the model. The 24 matched tumour-normal tissue samples from a subset of the participants were included in the training set and kept out of the testing set, as they were used to determine the concordance between methylation patterns between tumor versus cell-free DNA. Using the DMRs identified as concordant with the DMCs, we fitted a generalized linear model with elastic-net penalty function in the train set. We then used penalized logistic regression with five-fold cross validation to select the best predictive DMRs in the test set. The methylated regions were modeled as a single variable representing linear combinations of methylation markers in the test set, weighted by their regularized coefficients, termed cell-free Methylation Risk Score (cfMRS) (**Supplementary Table 2**). The model performance was evaluated by area under the receiver operating characteristics (AUC) curves. To compare the added value of model performance after including methylation markers, three models were assessed: (a) risk factors only, including age, sex, smoking status and BMI, (b) cfMRS only and (c) combined model with both risk factors and cfMRS. Identical model coefficients were applied to the test set for model evaluation.

#### Survival analysis and prediction of survival

Survival analysis was conducted using the methylation profiles from all 199 pancreatic cases. We filtered out methylation sites with less than 10 read counts across all pancreatic cancer patients. The overall survival time was defined as the time from date of pancreatic cancer diagnosis to the date of death or the last known date alive. We applied a two-step approach to select the prognostic methylation markers. We first used Cox proportional hazards model to assess the effects of each methylation site on pancreatic cancer survival adjusted for age and sex and we retained markers significant at FDR P-value of <0.05. In the second step, we fitted generalized linear models with L1 penalty in the methylation markers retained from above Cox proportional hazards model. We used nested cross-validation method to calculate the risk score predictions for all cases. In the outer level of cross-validation, samples are split into training and testing samples. Model parameters are tuned by cross-validation within training samples only.

We divided all cases into two groups based on the median predicted risk score from above. We estimated survival rates using the Kaplan-Meier method and calculated median survival times. The Log-rank test was used to examine the differences in survival times by median risk score group.

#### External validation of survival prediction

We integrated data from two projects for external validation. DNA methylation data and clinical data (n=185) of the TCGA-PAAD project were downloaded from the Cancer Genome Atlas (https://portal.gdc.cancer.gov/). Similarly, DNA methylation data and clinical data (n⍰=461) of the ICGC-PACA-AU project were downloaded from the International Cancer Genome Consortium (ICGC) database (https://www.icgc.org). DNA samples for methylation analyses were extracted from tumour tissue specimens. The methylation data were generated based on the Illumina Infinium Human Methylation 450 BeadChip (450K array). After excluding cancer cases without methylation array data, and missing survival time, a total of 283 cases were used in the validation analysis. We identified the methylation sites in the Methylation Array that overlapped the methylation regions in our data. We used the weights for methylation markers estimated in our data to compute the risk score in the validation dataset and followed the same analysis steps as above.

##### Pathway analysis and network map

R package *annotatr*^*24*^ was used to annotate genomic regions and identify genes in the region of interest, and genes were mapped to the Gene Ontology (GO) pathway terms. We used *clusterProfiler*^*25*^ and g:*Profiler*^*26*^ R packages to perform overrepresentation analysis of GO pathway terms. We also developed co-methylation networks from DMRs using the Weighted Correlation Network Analysis (WGCNA) method^27^. Briefly, we used pairwise Pearson’s correlations between methylation values to establish a signed network. To ensure optimal network structure, a soft threshold of 8 was applied based on the scale-free topology criterion. Subsequently, we employed hierarchical clustering in conjunction with a topological overlap matrix (TOM)-based dissimilarity measure to create a dendrogram of the network. The branches of this dendrogram, identified using the dynamic tree cut function, represented individual modules. Module Eigengene representing its primary principal component was computed. Subsequently, the identified modules were visualized using *cytoscape*^*28*^.

## Data Availability

All data produced in the present study are available upon reasonable request to the corresponding author.

## Grant support/acknowledgements

The study is supported by the Canadian Cancer Society (CCS 704716) and Canadian Institute of Health Research (CIHR FDN 167273).

## Author contributions

RJH conceived and designed the study; RJH, SG, DDC, AB, PCZ, DC, and SYS were involved in the acquisition, analysis or interpretation of data. SB and YB conducted the statistical analyses. SB drafted the manuscript with critical input from RJH. All authors contributed to the interpretation of results and revision of the manuscript.

## Data availability

The cfMeDIP–seq next-generation sequencing data for patient samples that support the findings of this study are available upon request from the corresponding author to comply with institutional ethics regulation and patient consent requirement.

## Code availability

R scripts used to analyse the data are available at https://github.com/budsans/cfDNA_methylation_PCa

