## Supplemental Figures and Tables for "Plasma cell-free DNA methylome as early detection and prognostic marker for pancreatic cancer"

**Supplementary information**

**Supplementary Figure 1.** A genome-wide analysis of cell free DNA methylome in pancreatic cancer patients and healthy controls.

**Supplementary Figure 2.** Hazard ratios and 95% CI of pancreatic cancer survival in models containing methylation risk score and other clinical covariates.

**Supplementary Table 1.** Differentially methylated regions obtained from cfMeDIP-seq of pancreatic cancer patients (cases) and healthy donors (controls).

**Supplementary Table 2.** Beta coefficients of risk factors of pancreatic cancer in different models in Train set

**Supplementary Figure 1. A genome-wide analysis of cell free DNA methylome in pancreatic cancer patients and healthy controls. (A)** Volcano plot of methylated regions in the plasma cfDNA from pancreatic cancer cases and healthy controls. **(B)** Concordance between plasma cfDNA- and tumour-normal tissue derived methylated sites. **(C)** Permutation test to estimate the significance of concordance between plasma cfDNA- and tumour-normal tissue derived methylation sites.

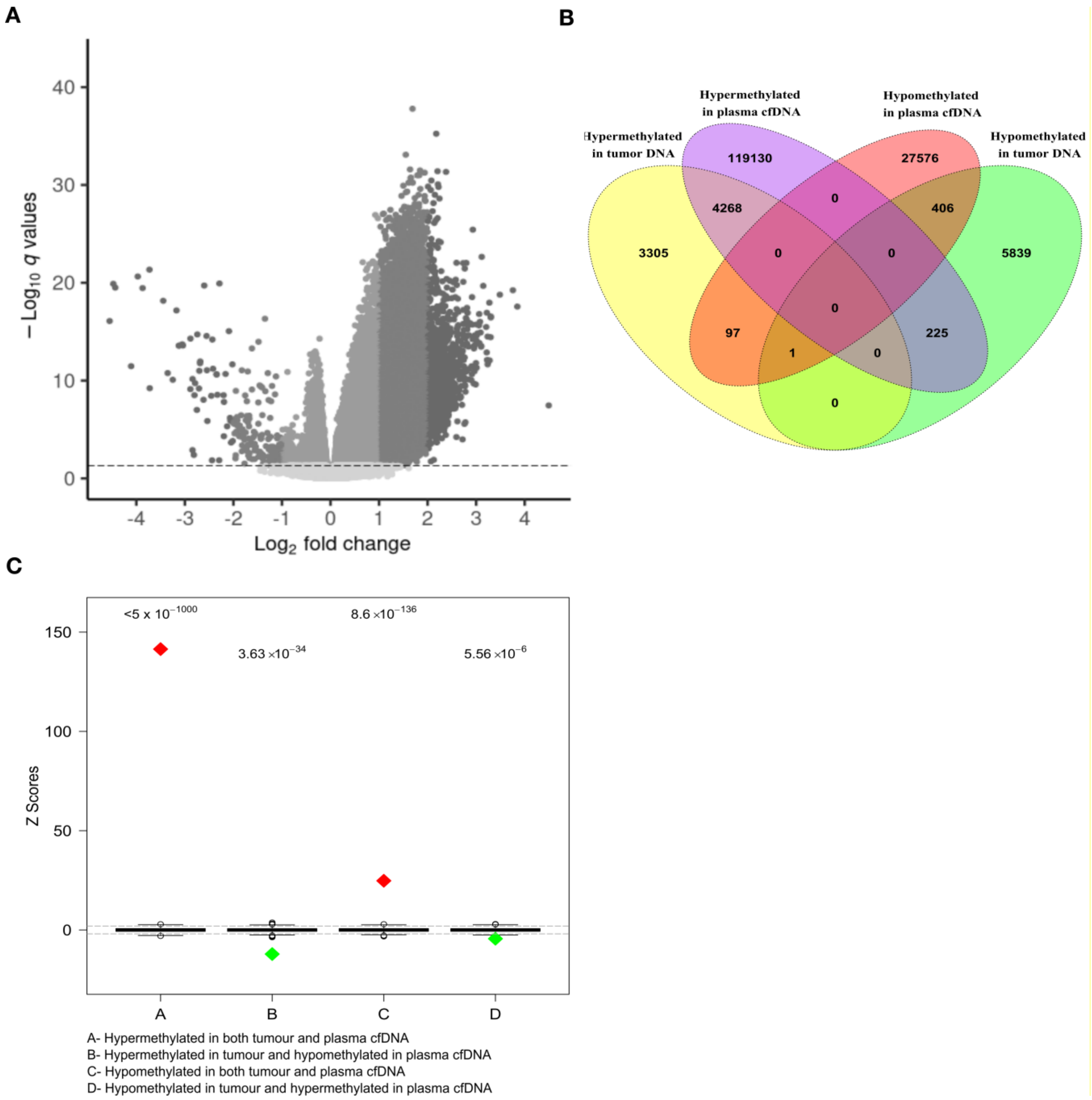

**Supplementary Figure 2.** Hazard ratios and 95% CI of pancreatic cancer survival in models containing methylation risk score and other clinical covariates.

| Model | Variables | Category | HR | 95%CI |
|---|---|---|---|---|
| Clinical variables | | | | |
| | Age (continuous) | | 1.02 | ( 1.01 - 1.04 ) |
| | Sex | Women | 1.00 | Ref. |
| | | Men | 1.00 | ( 0.72 - 1.38 ) |
| | Stage | I-II | 1.00 | Ref. |
| | | III-IV | 2.70 | ( 1.93 - 3.76 ) |
| cfMHS | | | | |
| | cfMHS category | Low risk (<= median) | 1.00 | Ref. |
| | | High risk (> median) | 5.69 | ( 3.96 - 8.17 ) |
| Clinical and cfMHS | | | | |
| | Age (continuous) | | 1.01 | ( 0.99 - 1.02 ) |
| | Sex | Women | 1.00 | Ref. |
| | | Men | 1.12 | ( 0.81 - 1.56 ) |
| | Stage | I-II | 1.00 | Ref. |
| | | III-IV | 1.68 | ( 1.19 - 2.38 ) |
| | cfMHS category | Low risk (<= median) | 1.00 | Ref. |
| | | High risk (> median) | 4.79 | ( 3.24 - 7.08 ) |

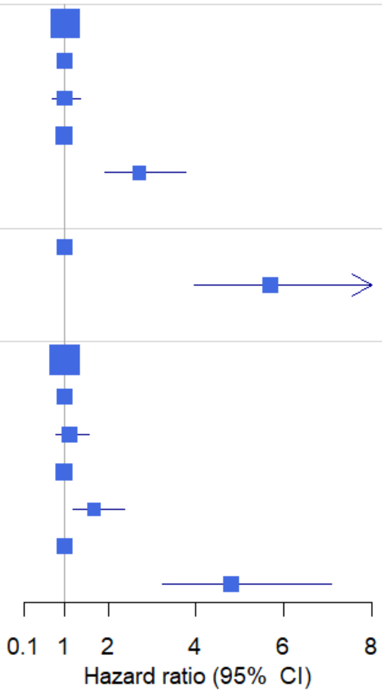


cfMHS, cell free methylation hazard score

**Supplementary Table 1.** Differentially methylated regions obtained from cfMeDIP-seq of pancreatic cancer patients (cases) and healthy donors (controls). P-values, Q-values and Log2 fold changes calculated using R package EdgeR; negative values show DMRs hypomethylated in cases and positive values show DMRs hypermethylated in cases, in comparison to controls.

| chrosome: start-end | logFC | PValue | FDR |
|---|---|---|---|
| chr1 : 110626501 - 110626800 | 1.66080552 | 2.2051E-10 | 1.3362E-07 |
| chr1 : 112058101 - 112058400 | 2.01085093 | 5.189E-32 | 5.1581E-27 |
| chr1 : 119530801 - 119531100 | 0.58655436 | 0.00040866 | 0.03052571 |
| chr1 : 156051001 - 156051300 | 1.09510161 | 3.1277E-15 | 5.1637E-12 |
| chr1 : 156405301 - 156405600 | 1.10088288 | 2.9469E-08 | 1.0892E-05 |
| chr1 : 16085701 - 16086000 | 1.06344538 | 9.9864E-11 | 6.5045E-08 |
| chr1 : 20140801 - 20141100 | -0.28897625 | 2.3842E-11 | 1.7687E-08 |
| chr1 : 221051701 - 221052000 | 1.08375541 | 3.8476E-05 | 0.00517517 |
| chr1 : 22333801 - 22334100 | -0.14878928 | 2.2587E-05 | 0.00336658 |
| chr1 : 228997201 - 228997500 | -0.31497676 | 1.6361E-07 | 4.9619E-05 |
| chr1 : 32404801 - 32405100 | -0.14075153 | 0.00023861 | 0.02084592 |
| chr1 : 3266101 - 3266400 | -0.27252429 | 3.434E-09 | 1.5916E-06 |
| chr1 : 36787801 - 36788100 | 0.46980079 | 5.2764E-09 | 2.3441E-06 |
| chr1 : 50881801 - 50882100 | 1.68707298 | 5.1889E-13 | 5.4492E-10 |
| chr1 : 91188301 - 91188600 | 0.73413498 | 0.00058893 | 0.039233 |
| chr10 : 12891301 - 12891600 | 0.57796819 | 0.00038601 | 0.02932913 |
| chr10 : 128994901 - 128995200 | 0.80198341 | 9.6349E-10 | 5.0765E-07 |
| chr10 : 133110001 - 133110300 | 1.16584524 | 0.00079675 | 0.04799746 |
| chr10 : 133955401 - 133955700 | -0.19369816 | 0.00028288 | 0.02357271 |
| chr10 : 77155801 - 77156100 | 0.31726264 | 0.00035882 | 0.0278605 |
| chr10 : 99790801 - 99791100 | 1.41001567 | 8.5405E-07 | 0.00021127 |
| chr11 : 10480801 - 10481100 | 0.87525111 | 0.00034094 | 0.02688874 |
| chr11 : 123172801 - 123173100 | 0.51549334 | 3.8611E-05 | 0.00518958 |
| chr11 : 128419201 - 128419500 | 1.47881136 | 1.1591E-30 | 6.8222E-26 |
| chr11 : 18812701 - 18813000 | 1.68572874 | 4.2701E-11 | 3.0118E-08 |
| chr11 : 2890201 - 2890500 | 0.76026184 | 3.6954E-06 | 0.00074366 |
| chr11 : 31847701 - 31848000 | 1.07791053 | 2.8431E-08 | 1.0553E-05 |
| chr11 : 3236701 - 3237000 | -0.24366806 | 1.2679E-07 | 3.9643E-05 |
| chr11 : 32453101 - 32453400 | 0.50271008 | 6.2753E-06 | 0.0011642 |
| chr11 : 45180301 - 45180600 | 0.4507588 | 2.3089E-05 | 0.00342806 |
| chr11 : 47416501 - 47416800 | 1.85824942 | 1.1181E-17 | 3.0664E-14 |
| chr11 : 55640401 - 55640700 | 0.24544013 | 0.00063422 | 0.04127623 |
| chr11 : 76849201 - 76849500 | 0.36056129 | 2.9054E-05 | 0.00413039 |
| chr12 : 119596201 - 119596500 | -0.38718561 | 1.234E-05 | 0.00204833 |
| chr12 : 132962401 - 132962700 | -0.28294111 | 1.874E-07 | 5.5985E-05 |
| chr12 : 53317801 - 53318100 | -0.11537422 | 0.00013958 | 0.01401125 |
| chr12 : 58014601 - 58014900 | 0.97510572 | 0.00034721 | 0.0272344 |
| chr13 : 28674601 - 28674900 | 1.62813417 | 8.761E-06 | 0.00153891 |
| chr13 : 33591601 - 33591900 | 0.87845549 | 0.00015927 | 0.01545939 |

| | | | |
|---|---|---|---|
| chr13 : 61987801 - 61988100 | 1.58737974 | 2.0094E-20 | 9.8665E-17 |
| chr13 : 84453601 - 84453900 | 1.84437337 | 6.6205E-16 | 1.2605E-12 |
| chr14 : 105944401 - 105944700 | 0.92946658 | 6.8429E-09 | 2.957E-06 |
| chr14 : 37131001 - 37131300 | 1.06842014 | 1.2404E-05 | 0.00205697 |
| chr14 : 95826601 - 95826900 | 0.29841527 | 0.00024069 | 0.02097505 |
| chr15 : 31775701 - 31776000 | 0.78175048 | 0.00048076 | 0.03418185 |
| chr15 : 34050601 - 34050900 | -0.2941846 | 7.2111E-08 | 2.4072E-05 |
| chr15 : 74421001 - 74421300 | 0.7426649 | 1.1811E-05 | 0.0019757 |
| chr15 : 75082201 - 75082500 | 0.8436332 | 0.00010676 | 0.01144845 |
| chr15 : 75084901 - 75085200 | 1.15379002 | 2.8465E-10 | 1.6864E-07 |
| chr15 : 83315101 - 83315400 | 1.13896889 | 2.3569E-13 | 2.656E-10 |
| chr16 : 1210501 - 1210800 | -0.33388709 | 1.9594E-07 | 5.8225E-05 |
| chr16 : 2090701 - 2091000 | -0.25028466 | 3.9071E-10 | 2.2473E-07 |
| chr16 : 2091001 - 2091300 | -0.26166783 | 0.00034397 | 0.02705335 |
| chr16 : 21294901 - 21295200 | 1.24223439 | 3.6894E-07 | 0.00010154 |
| chr16 : 30106501 - 30106800 | 0.5045177 | 2.1017E-06 | 0.00046026 |
| chr16 : 34659901 - 34660200 | 0.283287 | 0.00027059 | 0.02281907 |
| chr16 : 56703901 - 56704200 | 0.88686324 | 0.00012929 | 0.01322837 |
| chr16 : 56709901 - 56710200 | 0.63086561 | 0.00040738 | 0.03045886 |
| chr16 : 57831601 - 57831900 | -0.26907805 | 9.8864E-05 | 0.01080869 |
| chr16 : 71460001 - 71460300 | 0.76329615 | 0.00025589 | 0.02192649 |
| chr16 : 85660201 - 85660500 | 0.26148795 | 0.00046383 | 0.03333314 |
| chr16 : 86232301 - 86232600 | 1.64745425 | 3.0804E-23 | 2.7837E-19 |
| chr16 : 86530201 - 86530500 | 0.78871986 | 5.4867E-05 | 0.00684994 |
| chr16 : 86549101 - 86549400 | 1.27293856 | 3.1652E-08 | 1.1606E-05 |
| chr16 : 89268901 - 89269200 | 0.24553796 | 0.00010338 | 0.01117862 |
| chr16 : 9107101 - 9107400 | 0.53421076 | 6.2008E-05 | 0.00753971 |
| chr17 : 14733901 - 14734200 | -0.28862585 | 3.6293E-05 | 0.00494079 |
| chr17 : 18891901 - 18892200 | -0.19851786 | 0.00043878 | 0.03207075 |
| chr17 : 32907601 - 32907900 | 1.10742099 | 0.00038702 | 0.02938722 |
| chr17 : 33822601 - 33822900 | 0.85841932 | 1.4369E-08 | 5.7378E-06 |
| chr17 : 40260001 - 40260300 | 0.35545022 | 7.7478E-05 | 0.00896225 |
| chr17 : 42835501 - 42835800 | 0.60990413 | 6.8669E-07 | 0.00017466 |
| chr17 : 42835801 - 42836100 | 0.84126075 | 2.7475E-09 | 1.3036E-06 |
| chr17 : 46669501 - 46669800 | 0.88571633 | 3.4983E-07 | 9.6949E-05 |
| chr17 : 47073301 - 47073600 | 1.3114675 | 7.5818E-05 | 0.00881407 |
| chr17 : 47968201 - 47968500 | 0.31085962 | 0.0004066 | 0.03041819 |
| chr17 : 73749901 - 73750200 | 1.00945628 | 1.1525E-07 | 3.6449E-05 |
| chr17 : 75284101 - 75284400 | -0.26487629 | 1.3825E-05 | 0.00225024 |
| chr17 : 77179201 - 77179500 | 1.21936585 | 0.00066878 | 0.04274295 |
| chr17 : 77229601 - 77229900 | -0.24069827 | 3.5979E-05 | 0.00490651 |
| chr17 : 77777701 - 77778000 | 1.25980617 | 8.3661E-05 | 0.00950468 |
| chr17 : 79136401 - 79136700 | -0.18866136 | 3.6372E-06 | 0.0007337 |
| chr17 : 81014701 - 81015000 | -0.26933528 | 6.2656E-07 | 0.00016144 |
| chr18 : 44787901 - 44788200 | 0.38360799 | 0.0001685 | 0.0161203 |

| | | | |
|---|---|---|---|
| chr18 : 5890801 - 5891100 | 1.20333406 | 0.00079508 | 0.04793515 |
| chr18 : 9434401 - 9434700 | 0.25216887 | 1.4776E-05 | 0.00237662 |
| chr19 : 10023601 - 10023900 | 0.55349786 | 0.00012173 | 0.01264582 |
| chr19 : 1086601 - 1086900 | 0.34004997 | 3.418E-09 | 1.5851E-06 |
| chr19 : 1524001 - 1524300 | 1.38397447 | 8.968E-09 | 3.7637E-06 |
| chr19 : 16022701 - 16023000 | 1.4489286 | 8.3708E-19 | 2.9117E-15 |
| chr19 : 17008501 - 17008800 | 0.53437258 | 0.0003535 | 0.02757218 |
| chr19 : 18075001 - 18075300 | -0.24940325 | 4.5981E-11 | 3.2191E-08 |
| chr19 : 3428401 - 3428700 | 0.48984205 | 4.1823E-08 | 1.4887E-05 |
| chr19 : 35068201 - 35068500 | 0.26645753 | 2.5828E-06 | 0.00054868 |
| chr19 : 36049201 - 36049500 | 0.80317571 | 9.9001E-07 | 0.00024041 |
| chr19 : 36335101 - 36335400 | 0.61713505 | 9.3223E-07 | 0.00022812 |
| chr19 : 36643201 - 36643500 | 0.34284559 | 9.6867E-07 | 0.00023594 |
| chr19 : 3880801 - 3881100 | 1.01791391 | 0.0004508 | 0.03267235 |
| chr19 : 39998101 - 39998400 | 0.5505481 | 0.0007941 | 0.04789489 |
| chr19 : 4543501 - 4543800 | -0.30913569 | 3.7767E-05 | 0.00509979 |
| chr19 : 4543801 - 4544100 | -0.34578429 | 2.2699E-05 | 0.00338007 |
| chr19 : 49340401 - 49340700 | 1.06653196 | 7.7556E-06 | 0.00138912 |
| chr19 : 50393101 - 50393400 | 0.3793651 | 5.7172E-05 | 0.00707358 |
| chr19 : 53561101 - 53561400 | 0.36486749 | 8.78E-06 | 0.00154161 |
| chr19 : 54024601 - 54024900 | 0.73693299 | 7.414E-07 | 0.0001866 |
| chr19 : 55964101 - 55964400 | 0.40112013 | 0.00064259 | 0.04162982 |
| chr19 : 57149401 - 57149700 | 1.25808784 | 8.0926E-08 | 2.6678E-05 |
| chr19 : 57614701 - 57615000 | 0.56664321 | 0.00025865 | 0.02209746 |
| chr19 : 58513201 - 58513500 | 0.49655538 | 0.00080798 | 0.0484443 |
| chr19 : 947401 - 947700 | 0.30889925 | 6.1472E-09 | 2.6853E-06 |
| chr2 : 10054801 - 10055100 | -0.13178123 | 0.00029119 | 0.02405569 |
| chr2 : 109746301 - 109746600 | 0.58645077 | 0.0002812 | 0.02346258 |
| chr2 : 109746601 - 109746900 | 0.46606988 | 0.00037286 | 0.02862687 |
| chr2 : 112124101 - 112124400 | 0.293308 | 1.5431E-05 | 0.00246404 |
| chr2 : 113933701 - 113934000 | 0.70010215 | 0.00027182 | 0.02289307 |
| chr2 : 130763701 - 130764000 | 0.39631166 | 0.0002773 | 0.02322484 |
| chr2 : 1608901 - 1609200 | 0.52481015 | 1.7461E-07 | 5.2568E-05 |
| chr2 : 161504701 - 161505000 | -0.24665346 | 8.6259E-05 | 0.00973253 |
| chr2 : 171572701 - 171573000 | 0.77749648 | 0.00026196 | 0.02229871 |
| chr2 : 171679501 - 171679800 | 1.61567889 | 1.9149E-14 | 2.6962E-11 |
| chr2 : 171680101 - 171680400 | 0.93700551 | 0.00013448 | 0.01362849 |
| chr2 : 172952701 - 172953000 | 1.16882321 | 3.9244E-09 | 1.7961E-06 |
| chr2 : 172973701 - 172974000 | 0.50893246 | 0.00070228 | 0.04414918 |
| chr2 : 175199401 - 175199700 | 1.38901781 | 7.046E-06 | 0.00128225 |
| chr2 : 177027301 - 177027600 | 0.94668405 | 2.3451E-10 | 1.4121E-07 |
| chr2 : 214582201 - 214582500 | 0.32567957 | 1.7323E-06 | 0.00039033 |
| chr2 : 220361401 - 220361700 | 1.48062867 | 8.4878E-15 | 1.2844E-11 |
| chr2 : 227660701 - 227661000 | -0.29831008 | 6.8015E-10 | 3.7054E-07 |
| chr2 : 240493501 - 240493800 | -0.17542791 | 0.00036907 | 0.02841843 |

| | | | |
|---|---|---|---|
| chr2 : 35616901 - 35617200 | -0.22273767 | 2.3864E-09 | 1.1491E-06 |
| chr2 : 45158701 - 45159000 | 0.99832226 | 1.8215E-09 | 9.0058E-07 |
| chr2 : 69002101 - 69002400 | 2.14936645 | 2.996E-35 | 1.0309E-29 |
| chr2 : 71128801 - 71129100 | 0.4771095 | 1.5738E-06 | 0.00035923 |
| chr2 : 72371101 - 72371400 | 1.21869994 | 2.5729E-18 | 8.0484E-15 |
| chr2 : 85106701 - 85107000 | 0.45916802 | 2.9683E-05 | 0.00420243 |
| chr2 : 88849201 - 88849500 | 0.44620849 | 0.00010831 | 0.01157565 |
| chr20 : 19984201 - 19984500 | 0.67313251 | 6.8459E-10 | 3.7254E-07 |
| chr20 : 20346001 - 20346300 | 1.22770807 | 5.7466E-14 | 7.3178E-11 |
| chr20 : 22656001 - 22656300 | 0.22191747 | 0.00057054 | 0.03839465 |
| chr20 : 23636101 - 23636400 | 1.05740199 | 9.5565E-17 | 2.1579E-13 |
| chr20 : 291001 - 291300 | 0.80621804 | 0.00017264 | 0.01641857 |
| chr20 : 34190101 - 34190400 | 0.2150631 | 0.0002351 | 0.02061995 |
| chr20 : 43344001 - 43344300 | -0.24190089 | 7.9913E-06 | 0.00142451 |
| chr20 : 44640901 - 44641200 | 0.43913859 | 0.00019724 | 0.01813521 |
| chr20 : 47442901 - 47443200 | 1.28283593 | 1.0653E-15 | 1.9418E-12 |
| chr20 : 58406701 - 58407000 | -0.18191205 | 4.1752E-08 | 1.4865E-05 |
| chr20 : 60223201 - 60223500 | -0.21476904 | 0.0005977 | 0.03962754 |
| chr20 : 60951601 - 60951900 | -0.24808782 | 1.7464E-06 | 0.00039294 |
| chr20 : 61703401 - 61703700 | 1.05869922 | 1.234E-05 | 0.00204841 |
| chr20 : 62198701 - 62199000 | 0.85424925 | 2.5471E-17 | 6.4958E-14 |
| chr21 : 37914601 - 37914900 | 0.51293095 | 1.8483E-08 | 7.1812E-06 |
| chr21 : 38070301 - 38070600 | 0.75418545 | 0.00024888 | 0.02149942 |
| chr21 : 46125901 - 46126200 | 0.31119758 | 9.88E-06 | 0.00170212 |
| chr21 : 46572301 - 46572600 | 1.16139134 | 6.5939E-24 | 6.924E-20 |
| chr21 : 46836001 - 46836300 | -0.25138816 | 0.00016712 | 0.0160241 |
| chr22 : 20225101 - 20225400 | -0.40751935 | 1.1358E-07 | 3.5987E-05 |
| chr22 : 37420201 - 37420500 | -0.40872463 | 3.2139E-12 | 2.8596E-09 |
| chr22 : 38220601 - 38220900 | 0.8477878 | 2.0605E-07 | 6.0849E-05 |
| chr22 : 39160201 - 39160500 | -0.22732519 | 2.5793E-09 | 1.2317E-06 |
| chr22 : 46496101 - 46496400 | -0.19036585 | 0.00015082 | 0.0148462 |
| chr22 : 48886501 - 48886800 | 0.62362331 | 0.00039753 | 0.02994568 |
| chr22 : 51169801 - 51170100 | -0.20174165 | 0.00035856 | 0.02784641 |
| chr3 : 10749901 - 10750200 | 0.52211461 | 6.7302E-05 | 0.00804064 |
| chr3 : 115510201 - 115510500 | -0.27124862 | 0.00024856 | 0.02148054 |
| chr3 : 127795801 - 127796100 | 1.3714279 | 6.7162E-10 | 3.6642E-07 |
| chr3 : 129324301 - 129324600 | 1.46013083 | 7.9376E-09 | 3.374E-06 |
| chr3 : 133393501 - 133393800 | 0.72071917 | 2.4371E-10 | 1.4629E-07 |
| chr3 : 174126301 - 174126600 | -0.20365888 | 6.9907E-05 | 0.00827632 |
| chr3 : 197392801 - 197393100 | -0.26731139 | 9.8515E-05 | 0.01078089 |
| chr3 : 35706001 - 35706300 | 0.83087962 | 1.9417E-09 | 9.5339E-07 |
| chr3 : 38157901 - 38158200 | -0.37913627 | 2.3671E-08 | 8.9627E-06 |
| chr3 : 49843201 - 49843500 | 1.34578834 | 1.5023E-10 | 9.4304E-08 |
| chr3 : 52861201 - 52861500 | -0.29868774 | 9.1485E-06 | 0.00159563 |
| chr4 : 15907501 - 15907800 | -0.2662533 | 0.00077006 | 0.04694569 |

| | | | |
|---|---|---|---|
| chr4 : 41748001 - 41748300 | 0.87888019 | 9.4377E-05 | 0.01043498 |
| chr4 : 42153601 - 42153900 | 1.29604215 | 0.00039686 | 0.02990955 |
| chr4 : 4863301 - 4863600 | 0.90306211 | 6.6277E-10 | 3.6199E-07 |
| chr5 : 100239901 - 100240200 | 1.33967782 | 4.4327E-07 | 0.00011928 |
| chr5 : 10562701 - 10563000 | 0.27168724 | 0.00079001 | 0.04773516 |
| chr5 : 140616301 - 140616600 | 0.4186463 | 0.00025353 | 0.02178414 |
| chr5 : 149546701 - 149547000 | 0.411851 | 8.566E-05 | 0.00968033 |
| chr5 : 50259901 - 50260200 | 0.83845866 | 0.00078608 | 0.04758382 |
| chr5 : 76941601 - 76941900 | 0.43034915 | 6.3893E-05 | 0.00771866 |
| chr6 : 10113601 - 10113900 | 0.70118635 | 2.8029E-05 | 0.00401207 |
| chr6 : 119558401 - 119558700 | 0.31835654 | 0.00022908 | 0.02023327 |
| chr6 : 27258901 - 27259200 | 0.69087222 | 6.3305E-06 | 0.00117292 |
| chr6 : 27259201 - 27259500 | 0.51186945 | 0.00040167 | 0.03015748 |
| chr6 : 30139801 - 30140100 | 1.99047731 | 3.0994E-15 | 5.1226E-12 |
| chr6 : 36930601 - 36930900 | 0.30097397 | 1.163E-05 | 0.00195002 |
| chr7 : 100882501 - 100882800 | 0.50074819 | 8.3143E-05 | 0.00946041 |
| chr7 : 101005801 - 101006100 | 0.61644976 | 2.2437E-08 | 8.5352E-06 |
| chr7 : 102066001 - 102066300 | 0.53568817 | 8.0467E-13 | 8.1033E-10 |
| chr7 : 1164601 - 1164900 | -0.29941734 | 4.405E-08 | 1.5579E-05 |
| chr7 : 1286701 - 1287000 | 0.99020973 | 0.00071477 | 0.0446718 |
| chr7 : 151107601 - 151107900 | 0.54943612 | 0.00013732 | 0.01384313 |
| chr7 : 153585601 - 153585900 | 0.58184121 | 0.00012082 | 0.01257531 |
| chr7 : 158909101 - 158909400 | -0.4428408 | 1.2858E-06 | 0.00030189 |
| chr7 : 30722101 - 30722400 | 0.99213454 | 0.0002703 | 0.02280028 |
| chr7 : 35494201 - 35494500 | 0.78910357 | 2.6382E-05 | 0.00381929 |
| chr7 : 409501 - 409800 | -0.18643702 | 0.0001378 | 0.01387844 |
| chr7 : 47611801 - 47612100 | -0.28272492 | 7.581E-10 | 4.088E-07 |
| chr7 : 4885201 - 4885500 | 0.48590674 | 0.00030205 | 0.02468983 |
| chr7 : 56297401 - 56297700 | 0.44106498 | 0.00042456 | 0.03135909 |
| chr7 : 73392601 - 73392900 | -0.17506442 | 0.00043292 | 0.03178105 |
| chr8 : 134072101 - 134072400 | 1.21255432 | 7.0362E-20 | 3.0617E-16 |
| chr8 : 143625901 - 143626200 | -0.44505855 | 1.2569E-14 | 1.8437E-11 |
| chr8 : 144238501 - 144238800 | -0.15095463 | 1.1292E-05 | 0.0019026 |
| chr8 : 22723201 - 22723500 | 0.7165331 | 8.1443E-11 | 5.4057E-08 |
| chr9 : 1045501 - 1045800 | 0.33166092 | 1.1418E-06 | 0.00027198 |
| chr9 : 110249401 - 110249700 | -0.28857542 | 0.00031119 | 0.02521119 |
| chr9 : 126135901 - 126136200 | 1.30677922 | 1.892E-09 | 9.3133E-07 |
| chr9 : 130516501 - 130516800 | -0.38150203 | 1.7807E-10 | 1.0996E-07 |
| chr9 : 135620701 - 135621000 | 0.95367396 | 2.3509E-12 | 2.1516E-09 |
| chr9 : 136428001 - 136428300 | -0.31682247 | 0.00016362 | 0.01577376 |
| chr9 : 37002301 - 37002600 | 0.65704559 | 2.1176E-05 | 0.00319314 |
| chr9 : 87656101 - 87656400 | 0.33385036 | 3.2062E-05 | 0.00447272 |
| chr9 : 976501 - 976800 | 0.96405474 | 0.00037181 | 0.02856915 |

**Supplementary Table 2.** Beta coefficients of risk factors of pancreatic cancer in different models in Train set

| Model | Variables | Estimate |
|---|---|---|
| Risk factor | | |
| | Age (year) | -0.01 |
| | Sex: Men | -0.32 |
| | Smoking Status: Yes | 0.17 |
| | Diabetes: Yes | 0.96 |
| | Family cancer history: Yes | 1.4 |
| | BMI (Kg/m2) | 0.08 |
| Methylation marker | | |
| | Methylation risk score | 17.4 |
| Risk factor and Methylation marker | | |
| | Age (year) | 0.04 |
| | Sex: Men | 3.82 |
| | Smoking Status: Yes | -0.83 |
| | Diabetes: Yes | 1.44 |
| | Family cancer history: Yes | -1.09 |
| | BMI (Kg/m2) | -0.76 |
| | Methylation risk score | 16.37 |
